# A bench-to-data analysis workflow for respiratory syncytial virus whole-genome sequencing with short and long-read approaches

**DOI:** 10.1101/2025.04.07.25325192

**Authors:** Adrián Gómez-Del Rosario, Adrián Muñoz-Barrera, Julia Alcoba-Florez, Diego García-Martínez de Artola, Nora Rodríguez-García, Jose Miguel Lorenzo-Salazar, Rafaela González-Montelongo, Carlos Flores, Laura Ciuffreda

## Abstract

Respiratory syncytial virus (RSV) is a leading cause of acute respiratory illness, especially in children, immunocompromised patients, and the elderly. Genomic surveillance of RSV enables detecting and monitoring of circulating lineages and the emergence of amino acid substitutions affecting transmission, severity, and treatment, including resistance to monoclonal antibodies (mAb). Most available RSV genomic surveillance protocols target specific regions of the viral genome, such as the *G* and *F* genes, or are designed for a particular sequencing technology. Here, we aimed to develop and validate a comprehensive workflow for whole-genome RSV sequencing adapted for short- and long-read sequencing data, covering all steps from sample processing to lineage classification. Viral RNA was collected from 176 nasopharyngeal swab samples from patients in the Canary Islands (Spain) over two seasons between October 2022 and February 2024. Molecular diagnosis was performed with RT-qPCR and genomic libraries were prepared in parallel with tiling amplicon protocols adapted for Illumina and Oxford Nanopore Technologies (ONT) sequencing approaches. An in-house bioinformatics pipeline built on Nextflow was also developed to enable deployment, ensure built-in dependency management, and provide adaptability to both sequencing technologies. We found a high concordance between Illumina and ONT sequencing results in terms of consensus sequence, lineage assignment, and lineage-defining substitutions. We also identified RSV sequences harboring substitutions associated with mAbs resistance and patient coinfections. Taken together, the developed resources and results allowed us to provide for the first time detailed genomic data of RSV transmissions within the epidemiological context in the Canary Islands.

## Introduction

Respiratory syncytial virus (RSV) has a major impact on public health as it is the leading cause of acute lower respiratory tract infections in infants and children under two years of age (Alvarez *et al*., 2013). There are two main antigenic subgroups of RSV (RSVA and RSVB), defined by the G glycoprotein, that co-circulate globally, with the predominance of one or the other depending on the different epidemic seasons. In the majority of worldwide RSV infections, patients are asymptomatic or have mild symptoms (Caserta *et al*., 2017; Mazur, Caballero and Nunes, 2024). However, some patients can develop severe illness that can lead to hospitalization (Vila et al., 2023). RSV can also severely affect adults, especially those with comorbidities, immunocompromised, or the elderly (Nguyen-Van-tam *et al*., 2022). In Spain, two out of every 100 children under two years of age are hospitalized every year, with an estimated annual cost to the National Health System of 50 million euros (Martinón-Torres *et al*., 2022; Gea-Izquierdo *et al*., 2023). On a global scale, the annual economic impact of RSV is estimated to be ~4.82 billion euros, considering only children under five years of age (Zhang *et al*., 2020). Substantial efforts have been made to develop an immunoprophylactic treatment strategy based on vaccination or neutralizing monoclonal antibodies (mAb). In March 2023, nirsevimab, a long-acting anti-RSV fusion protein mAb, was included in the immunization schedule of some Spanish regions, such as Galicia (Ares-Gómez *et al*., 2024) and the Canary Islands, and then into the rest of the country (Mestre-Ferrándiz *et al*., 2024). Although rare, nirsevimab-resistant RSV lineages have been reported (Ares-Gómez *et al*., 2024).

Genomic surveillance of RSV is crucial for the identification and monitoring of the circulating lineages as it provides insight into the emergence of amino acid substitutions associated with resistance to treatments and vaccines, as well as information on viral evolution and epidemiology. Few protocols have been established for this purpose, most of which target only specific RSV regions, such as the *G* and *F* genes. Here, we have adapted an RSV whole-genome tiling amplicon sequencing protocol, originally developed for Illumina sequencing (Davina-Nunez *et al*., 2023), to nanopore sequencing using Oxford Nanopore Technologies (ONT) and developed an in-house bioinformatic pipeline for the analysis of the genomic data generated by the two technologies (https://github.com/genomicsITER/nf-rsvpipeline). We have then tested and compared the results obtained by this novel workflow using clinical samples processed by both sequencing technologies in parallel. Our study represents one of the few RSV genomic surveillance efforts in Spain (Davina-Nunez *et al*., 2023; Rojo-Alba *et al*., 2024), and the first one in the Canary Islands.

## Materials and methods

### Sample collection from patients and diagnosis

A total of 176 nasopharyngeal swab samples were collected from 173 patients in Tenerife (Canary Islands, Spain) between October 2022 and February 2024, spanning two RSV seasons, and sent to the University Hospital Nuestra Señora de Candelaria (Santa Cruz de Tenerife, Spain) for molecular diagnosis. Samples were collected from patients who required hospital care (hospitalized, admitted to the emergency room or outpatients) with symptoms compatible with a respiratory viral infection or from immunosuppressed patients screened on admission. RNA was isolated from nasopharyngeal swabs using MagNA pure 96 DNA and Viral NA Small Volume kit (Roche Life Science [Basel, Switzerland]) and RSV infection was diagnosed by reverse transcription quantitative PCR (RT-qPCR) using two alternative kits (the Xpert^®^ Xpress CoV-2/Flu/RSV plus [Cepheid, Sunnyvale, CA, USA] and the Alinity m Resp-4-plex assay [Abbott Laboratories, Chicago, IL, USA]). The study was approved by the Research Ethics Committee of the Complejo Hospitalario Universitario de Canarias review board (CHUNSC_2024_48).

### Library preparation and sequencing

For Illumina sequencing, the protocol for library preparation was previously described (Davina-Nunez et al., 2023). Libraries were prepared using the Illumina CovidSeq Test (Illumina Inc., San Diego, CA, USA), quantified using Qubit™ dsDNA HS Assay Kit (Invitrogen, Eugene, CA, USA) and checked on a 4200 TapeStation (Agilent Technologies, Santa Clara, CA, USA). Sequencing was performed on an Illumina NextSeq 550 system using 1.4 pM as the final concentration, and spiked with 1% PhiX Control v3 (Illumina Inc.).

For ONT sequencing, RT-PCR was performed using the Midnight RT PCR Expansion kit (ONT, Oxford, UK), using the same primers employed for Illumina sequencing. Libraries were prepared using the Rapid Barcoding Kit 96 (ONT) and sequencing was performed using the R9.4.1 flow cells in a MinION Mk1B or a GridION X5 (ONT) for at least 48 h. Basecalling was performed using the MinKNOW v24.02.16 and the integrated basecaller Dorado v7.3.11 with the super-accuracy basecalling model.

### Bioinformatic pipeline for sequencing data analysis

Using Nextflow (Di Tommaso et al. 2017) and following nf-core (Ewels et al. 2020) guidelines and templates as framework, a full in-house bioinformatic pipeline was developed integrating state-of-the-art tools (**Table 1**) to generate consensus sequences of RSV viruses using Illumina or ONT sequencing data.

**Table 1.**
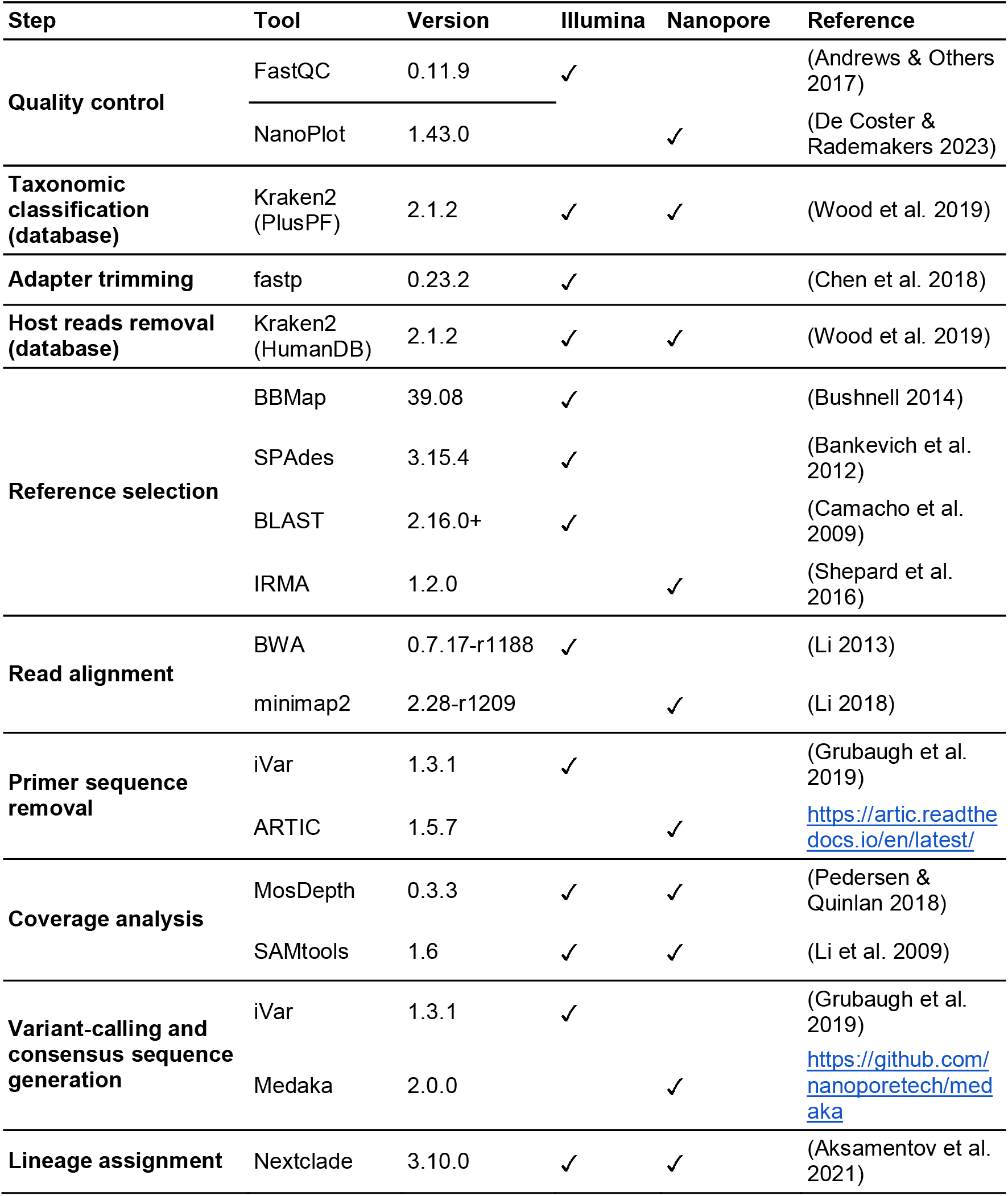
List of software tools used in the bioinformatic pipeline.

### Analysis of coinfections

To evaluate potential coinfections in some samples, an RT-qPCR using the Allplex™ Respiratory Panel 1 kit (Seegene, Seoul, South Korea) was performed. When low concordance for the lineage-defining mutations could not be explained by a loss of coverage in the sequence, alignment BAM files were manually inspected with Integrative Genome Viewer (IGV) v2.17.4 (Robinson *et al*., 2011). When discordance could not be explained by the presence of a RSVA/RSVB coinfection, ViralFlow (v1.1.1) (Da Silva *et al*., 2024) was used to confirm the presence of intra-host viral lineages of the same antigenic subgroup. A detailed analysis workflow is provided in **Figure 1**.

**Figure 1.**
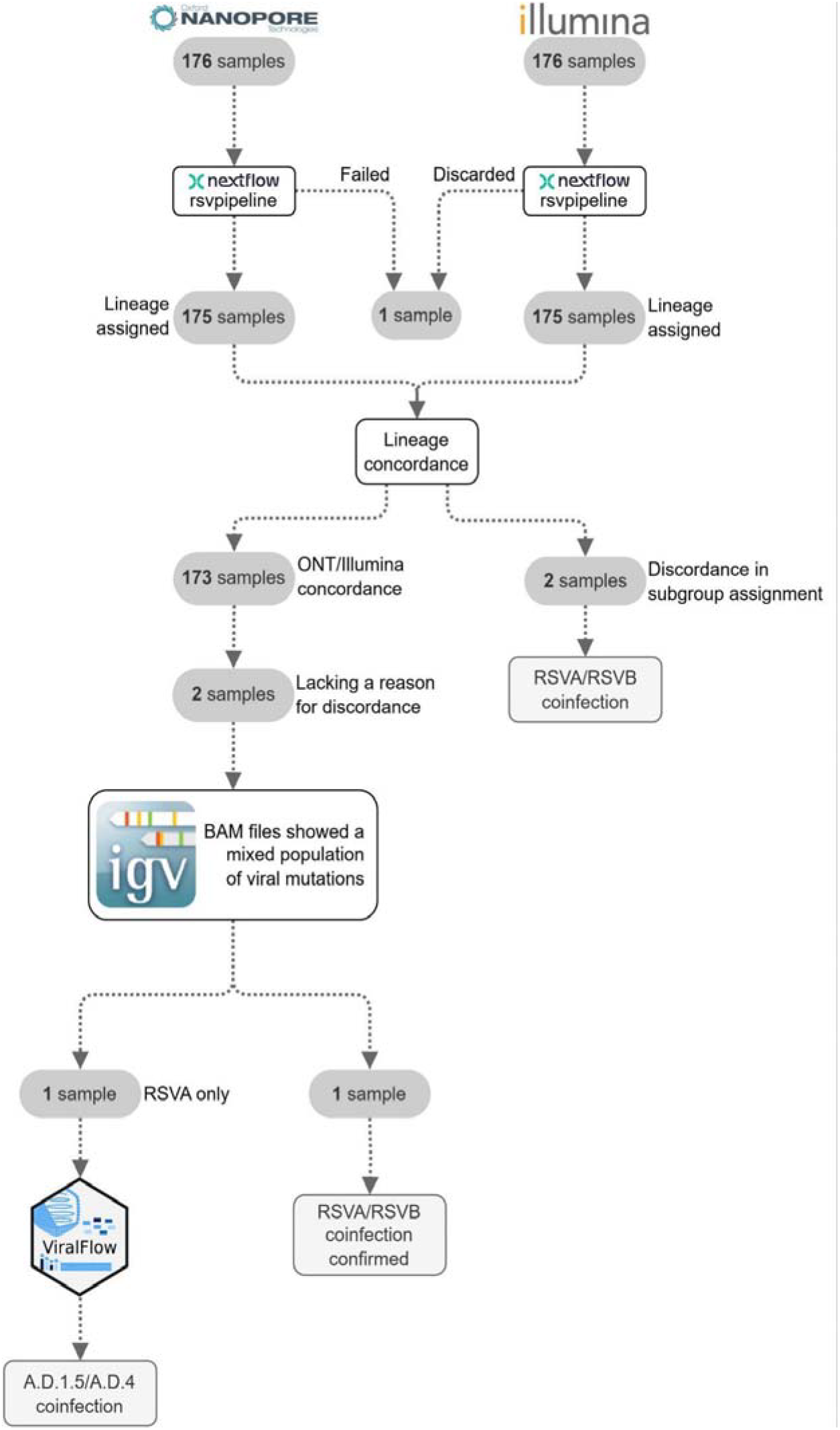
Flow chart of the analysis workflow.

### Amino acid substitutions associated with the efficacy of neutralizing monoclonal antibodies

For the analysis of the amino acid substitutions related to reduced efficacy to mAbs, we searched for amino acid changes in the nirsevimab binding site (site Ø of F protein, amino acid residues 62-69 and 196-212) and the palivizumab binding site (site II of F protein, amino acid residues 262-275), as previously described (Wilkins *et al*., 2023). Only samples that had both binding sites covered in the sequence were included in this analysis.

### Statistical analysis

To assess the differences between the Illumina and ONT sequencing strategies, we compared the genomic breadth of coverage, defined as the percentage of the consensus sequence covered, the lineage assigned to each consensus sequence by Nextclade, and the concordance in detecting the lineage-defining mutations following (Goya *et al*., 2024). For the analysis of the genomic breadth of coverage, a chi-square test was used. To compare the lineage-defining mutations, the concordance found by the two technologies was calculated for each mutation as the number of matches divided by the number of sequences assigned to the lineage. Data analysis was performed using R v4.3.2 (R Core Team, 2024). Statistical significance was established at *p*<0.05.

## Results

### Pipeline description

To generate the consensus sequences of RSV viruses using Illumina sequencing (**Figure 2A**), the developed pipeline starts with a quality control step performed using FastQC to assess sequence quality metrics. To confirm the presence of RSV in the samples, a taxonomic classification of reads is performed using Kraken2 with the PlusPF database. Adapter sequences are removed from the reads using fastp, and human-host reads are scrubbed by means of Kraken2 with the human-only database. The reference sequences *hRSV/A/England/397/2017* and *hRSV/B/Australia/VIC-RCH056/2019*, included in a multi-reference FASTA file, are used to identify the most appropriate RSV reference for downstream analysis. This is achieved by performing a multiple sequence alignment (MSA) of the reads using BBmap. If the MSA step fails, an alternative assembly strategy using SPAdes is performed, followed by a contig classification step using BLAST to assign the sample to the correct antigenic subgroup (RSVA/RSVB) and select the appropriate viral reference sequence. The remaining viral reads after scrubbing are mapped to the selected reference genome using BWA and primer adapters are trimmed using iVar. Genome-wide breadth coverage and specific coverage of the *G* and *F* genes are assessed using Mosdepth and SAMtools, ensuring adequate representation and depth across the viral genome. Mutations and consensus sequences are obtained by piping SAMtools pileup with the iVar derived consensus output, as described elsewhere (Croville *et al*., 2022). Finally, consensus sequences are submitted to the Nextclade web version for qualitative mutation calling, sequence quality assessments, and lineage assignment after phylogenetic placement.

**Figure 2.**
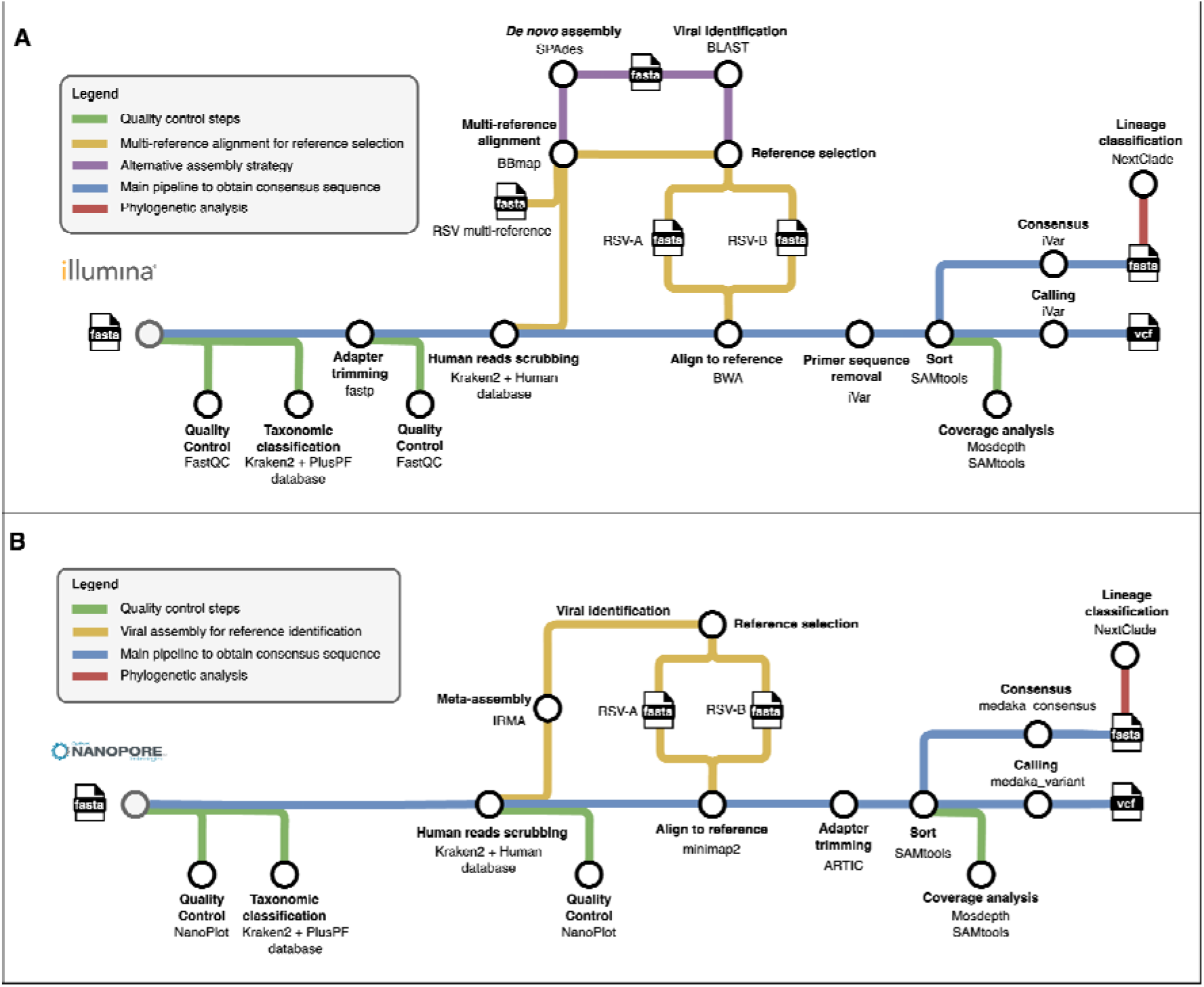
Bioinformatic pipeline to obtain the RSV consensus sequences from Illumina (**A**) or Oxford Nanopore Technologies (**B**) sequencing data.

Given the differences in read lengths and error profiles associated with Illumina and ONT raw data, specific tools and quality control steps are incorporated into the ONT pipeline to ensure accurate viral genome reconstruction (**Figure 2B**). Quality control is performed using NanoPlot. Then, taxonomic classification and host-read removal are performed using Kraken2 with the PlusPF database and the human-only database, respectively. The remaining reads are subjected to a meta-assembly step by means of IRMA, using the provided RSV-specific module, to select the most appropriate RSV reference genome for downstream analyses. The de-hosted viral reads are subsequently mapped to the selected RSV reference using minimap2, and adapter trimming is conducted with the align_trim tool from the ARTIC pipeline. Coverage analysis is performed using the same methods described for the Illumina pipeline. Finally, variant calling and consensus sequence generation are carried out using Medaka. Qualitative mutation calling, sequence quality assessments, and lineage assignment are performed using the Nextclade web version.

### Comparative analysis of RSV results by sequencing technology

A total of 176 samples were sequenced using the previously described Illumina and ONT protocols. The median number of reads obtained was 2.1M reads (range 0.81-5.0M reads) for Illumina and 95.2k reads (range 1-397k reads) for ONT. One sample failed to complete the pipeline due to the low number of reads obtained by ONT and was discarded from the comparative analyses. The average depth of coverage was 3,713X (standard deviation [SD]=3,892) for Illumina sequencing and 2,430X (SD=2,258) for ONT, with both technologies showing similar results in amplicon coverage (**Figure S1**). Consensus sequences covering at least 80% of the genome were obtained for 134 of the samples sequenced by Illumina (76.6%) and 172 of the samples sequenced by ONT (98.3%) (Chi-square test, p=2.44×10-9). Notably, at least 80% of the RSV genome of all samples with a cycle threshold (Ct) ≤25 were covered by both technologies (**Figure S2, Figure S3**). Lineage classification was concordant in 98.8% of the samples. Only two samples showed misclassification of RSVA/RSVB (**Table S1**) and were excluded from further analysis. Despite the high concordance, there were differences in the observed lineage-defining mutations (**Figure 3**). The concordance between the two technologies was 100% for all lineage-defining mutations in lineages

**Figure 3.**
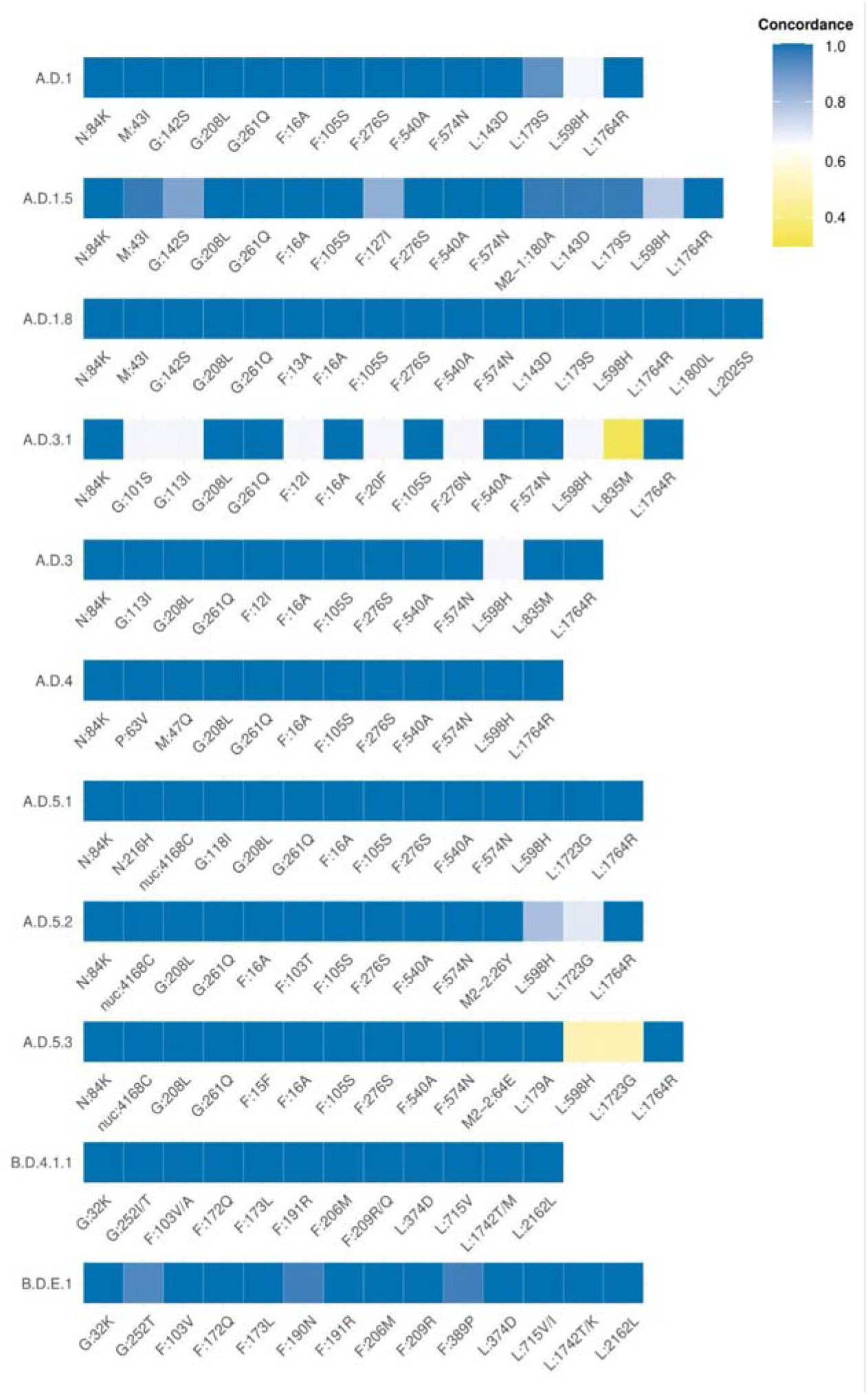
Heatmap of the Illumina and ONT concordance of the lineage-defining mutations, based on Goya et al. (2024).

A.D.1.8 (n=1), A.D.4 (n=11), A.D.5.1 (n=4), and B.D.4.1.1 (n=5). In the remaining lineages, concordance was also 100% for all mutations except for the following: one mutation for A.D.3 (n=3), two mutations in A.D.1 (n=12), A.D.5.2 (n=20), and A.D.5.3 (n=2), four mutations in B.D.E.1 (n=81), six mutations in A.D.1.5 (n=31), and seven mutations in A.D.3.1 (n=3). Note that the apparent accumulation of discordance in A.D.3, A.D.3.1, and A.D.5.3 is likely due to the low number of sequences assigned to these lineages (**Table S1**). Notably, a few previously undescribed amino acid substitutions were detected by the two technologies at the same position: the lineage-defining mutations in the B.D.4.1.1 (L:1742M, F:103A, and G:252T) and B.D.E.1 (L:1742K and L:715I) lineages.

For two samples, assigned to A.D.1.5 and A.D.5.2, the discordance in one mutation could not be explained by low coverage or the presence of an undetermined nucleotide in the consensus sequence, requiring further investigation. A detailed analysis of the read alignments against the RSV reference genome was performed in these instances, and the presence of different variants in the same locus was observed. One of the two samples was confirmed as RSVA/RSVB coinfection, despite it was previously classified exclusively as RSVA based on the data provided by the two technologies. As the other sample only showed evidence for RSVA, we re-analyzed the data using ViralFlow to check the presence of intra-host viral variant. ViralFlow provided additional support for a potential coinfection of two RSVA A.D.-like sublineages (A.D.1.5 and A.D.4) in this sample (**Figure 1)**.

### Epidemiological analysis and resistance to monoclonal antibodies

The newly developed bioinformatic pipeline and the results presented here provided detailed genomic data from RSV infections, allowing us to study for the first time the RSV epidemiological scenario in the Canary Islands. Among patients, 83 (47.98%) were male and 90 (52.02%) were female. Most patients were children up to two years of age (48.56%) and elderly over 60 (38.15%). Twenty-seven (15.6%) cases required hospitalization (**Table S2**). Defining reinfection as the occurrence of two samples diagnosed as RSV-positive collected within a maximum interval of 30 days, two cases of reinfection were observed among the 173 patients. Out of the 176 samples collected for this study, in the 2022-2023 season (n=120), RSVB lineages predominated with 60% of RSV infections attributed to this antigenic subgroup, most of them assigned to B.D.E.1 (**Figure 4A**). In contrast, during the 2023-2024 season (n=56), RSVA lineages predominated, representing 73.2% of cases, with the A.D.1.5 lineage being the most common (**Figure 4B**).

**Figure 4.**
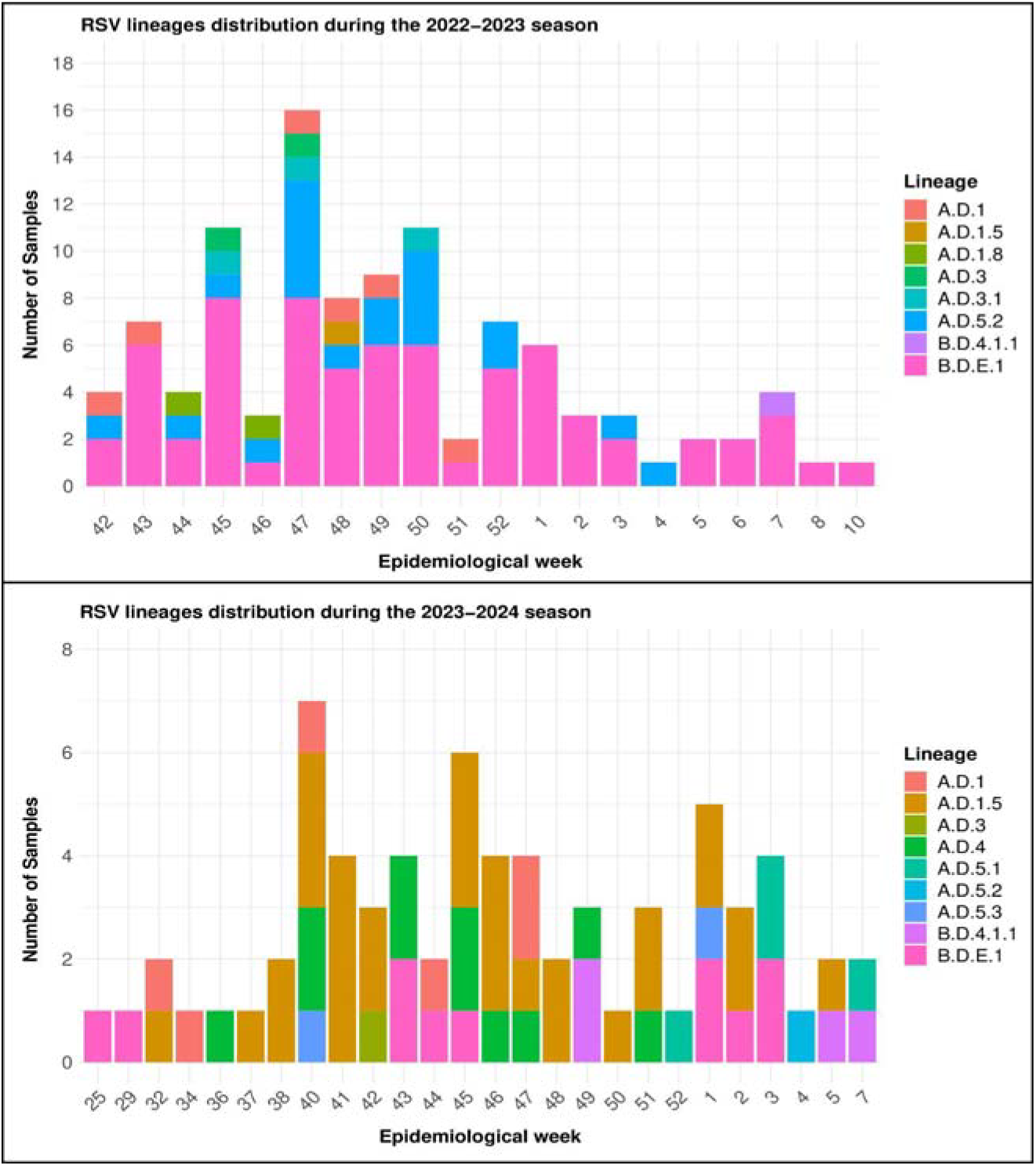
Nextclade lineage assignment series of RSV sequences from the 2022-2023 (**A**) and 2023-2024 (**B**) seasons.

To assess the occurrence of amino acid substitutions associated with potential resistance to mAb, we examined the binding sites palivizumab and nirsevimab. The target region of these mAbs was covered in the consensus sequence of 85 out of 87 RSVA samples. The following two amino acid changes were detected in the nirsevimab binding site: F:N63S in 12 of them (11 in A.D.4 and one in A.D.1.5), and F:I206T in one A.D.3.1. The target region of the mAbs was covered in the consensus sequence of 85 out of 86 RSVB samples, with four substitutions identified in the nirsevimab binding site: 84 carried the F:I206M and F:Q209R substitutions (80 B.D.E.1 and four B.D.4.1.1), 81 also carried the F:S211N substitution (80 B.D.E.1 and one B.D.4.1.1), and one B.D.E.1 also carried the F:K68N amino acid change. In addition, one B.D.E.1 lineage also carried the F:K272T substitution in the palivizumab binding site.

## Discussion

Genomic surveillance of RSV is of utmost importance for public health, especially for monitoring the potential emergence of nirsevimab-resistant amino acid substitutions of RSV (Wilkins *et al*., 2023). Here, we successfully adapted and validated a novel RSV genomic surveillance workflow for viral whole-genome sequencing of clinical samples. This workflow consists of a tiling amplicon sequencing protocol for short or long-read sequencing technologies and an in-house bioinformatic pipeline for the analysis of genomic data. Our results demonstrate that the workflow performs equally for both sequencing alternatives, successfully generating consensus sequences and highly concordant lineage classifications in clinical samples from two seasons in the Canary Islands. To date, most available workflows for genomic surveillance of RSV have been optimized for a single sequencing technology, either ONT or Illumina (Wang *et al*., 2022; Piñana *et al*., 2024; Talts *et al*., 2024; Madi *et al*., 2024), while only a few have been designed for both sequencing technologies (Dosbaa *et al*., 2024; Dong *et al*., 2025). To the best of our knowledge, this is the first bench-to-data analysis workflow developed for RSV whole-genome sequencing that has also made a direct technological comparison while validating the results with clinical samples. It also constitutes the first genomic assessment of the 2022-2023 and 2023-2024 epidemiological seasons of RSV in the Canary Islands and one of the few that have been performed to date in Spain (Davina-Nunez *et al*., 2023; Rojo-Alba *et al*., 2024).

Two of the 176 analyzed clinical samples were classified into different antigenic subgroups by the two technologies, suggesting the presence of coinfections in these samples and prompting further analyses including confirmatory RT-qPCR experiments. Based on the high concordance, the two sequencing technologies were on par to detect the lineage-defining mutations in clinical samples. However, some discordances were observed, especially in lineages A.D.1.5, A.D.5.2, and B.D.E.1. For Illumina sequencing, these discrepancies were primarily attributed to a loss of coverage in specific regions, leading to unassigned nucleotides in the consensus sequences. The observed difference between ONT and Illumina in the proportion of consensus sequences covering at least 80% of the genome may also be attributed to this loss of coverage. For ONT sequencing, discordance was due to frameshift errors in some of the consensus sequences. In two samples, none of these factors fully explained the discordance, and further analysis revealed the presence of a mixed viral population. Notably, one of the samples with RSVA/RSVB coinfections was obtained from a child with a T-cell lymphoblastic lymphoma who re-tested positive for RSVB only one month after the initial sample was collected. Even though RSVA/RSVB coinfections have been rarely reported so far (Yu et al. 2021), we have observed them in 3 out of 175 samples with assigned lineage analyzed, showing a prevalence of roughly 2% (1.7 every 100 cases) for the studied period. Our findings underscore the importance of continuous monitoring for coinfections in RSV genomic surveillance, particularly in immunocompromised patients, where the RSV burden is high (Gaillet *et al*., 2025; Ross *et al*., 2025). Additionally, they highlight how the sequencing with two technologies applied in parallel can enhance the detection and characterization of such cases.

We had particular interest in identifying the presence of amino acid changes present in the antigenic sites of two mAbs, nirsevimab (antigenic site Ø) and palivizumab (antigenic site II), to predict potential resistance to these treatments arising in the community. In the RSVA samples, we detected two substitutions in the nirsevimab binding site, F:N63S and F:I206T. These amino acid substitutions have been previously observed in RSV sequences and do not alter susceptibility to nirsevimab based on neutralization assays (Fourati, Zhu 2018, Bin Lu 2019, Lin 2022). In RSVB samples, four amino acid changes were identified in the nirsevimab binding site Ø; F:K68N, F:I206M, F:Q209R, and F:S211N, along with one substitution, F:K272T, in the palivizumab binding site II. According to some studies, the F:K68N substitution has been classified as a nirsevimab neutralization escape substitution (Wilkins *et al*., 2023; Rios-Guzman *et al*., 2024). In the present study, this substitution was observed in a sample collected during the first epidemiological week of 2023, before the introduction of nirsevimab in the Spanish immunization schedule. Despite the F:I206M, F:Q209R, and F:S211N substitutions have shown a high prevalence in RSVB samples, the actual evidence does not support a significant reduction in the nirsevimab efficacy associated with these amino acid changes (Fourati *et al*., 2024; Rios-Guzman *et al*., 2024). Only one sample assigned to the B.D.E.1 lineage was found to carry the F:K272T substitution, which is associated with palivizumab resistance (Zhu *et al*., 2012; Hashimoto and Hosoya, 2017; Wilkins *et al*., 2023).

This study has several limitations. First, the sample collection was limited to patients who required hospital care, therefore, data might not be representative of RSV transmission in the community. Second, some mutations may have been missed in some samples due to amplicon dropout, especially those with high Ct values. This issue could be mitigated in the future by adopting a recently published modified version of the tiling amplicon protocol (https://www.protocols.io/view/whole-genome-amplification-of-respiratory-syncytia-eq2lyjzbrlx9/v3). Third, the lack of information on whether patients received mAbs limited our ability to analyze RSV sequences in the context of breakthrough infections. However, our study also has strengths, as each sample was processed in parallel using the two sequencing technologies, enabling the detection of mixed RSV populations, which was validated through multiple approaches.

In conclusion, we developed a novel workflow for RSV genomic sequencing and validated it in clinical samples in a real-world setting. Our findings support a high performance with both short- and long-read sequencing technologies.

## Data Availability

Sequences in FASTA format have been deposited in GISAID. The code is publicly available and maintained within a GitHub repository (https://github.com/genomicsITER/nf-rsvpipeline).

## Data availability

The code is publicly available and maintained within a GitHub repository (https://github.com/genomicsITER/nf-rsvpipeline).

