## Supplementary material for "A bench-to-data analysis workflow for respiratory syncytial virus whole-genome sequencing with short and long-read approaches"

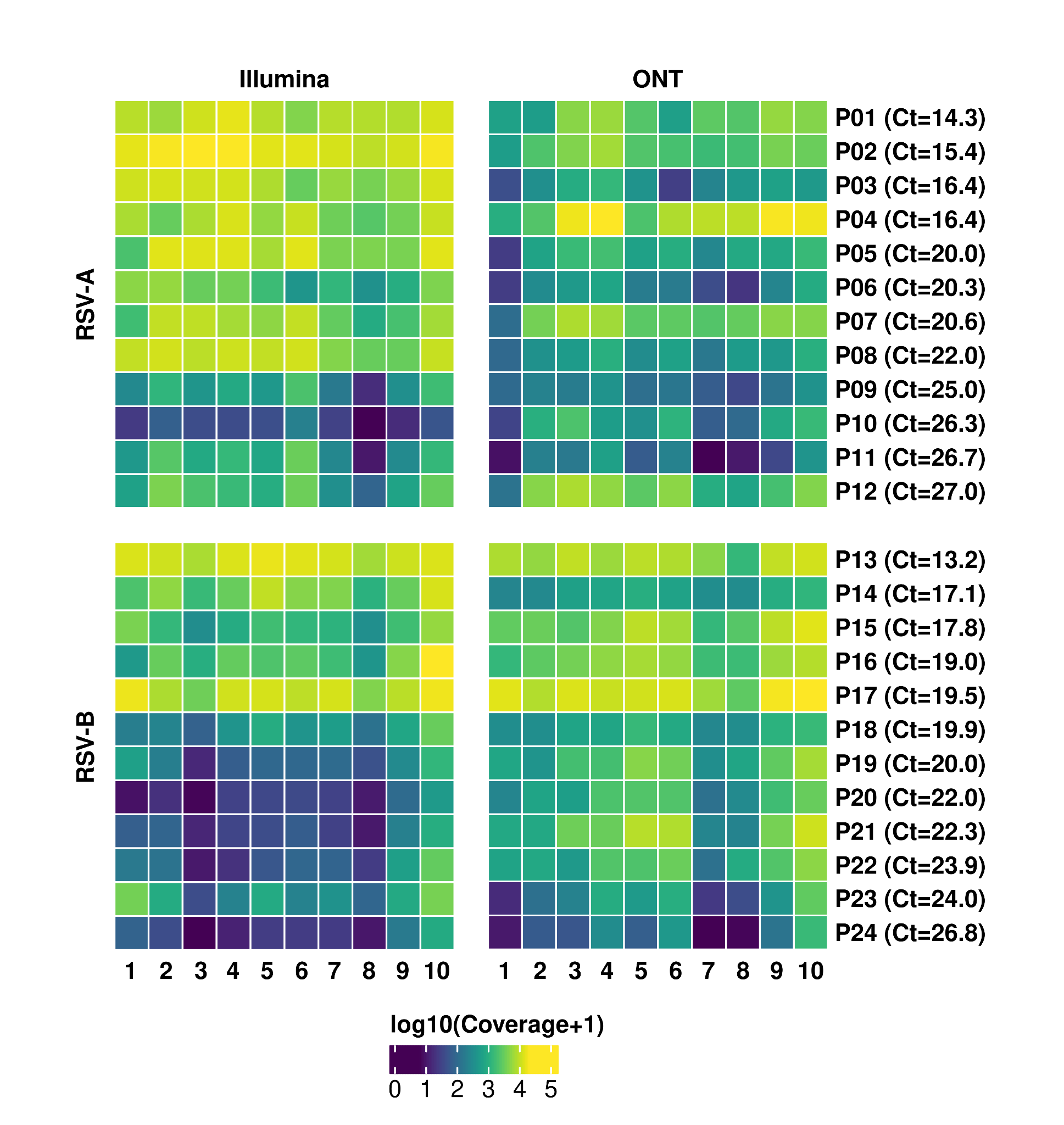

**Figure S1.** Heatmaps of median amplicon coverage across samples sequenced using Illumina (left) and ONT (right) for samples classified as RSV-A (top) and RSV-B (bottom). Labels in the horizontal axes represent the tiling amplicons used for each antigenic group. Samples are represented by P1 to P24. Ct values correspond to those observed qPCR cycle thresholds.

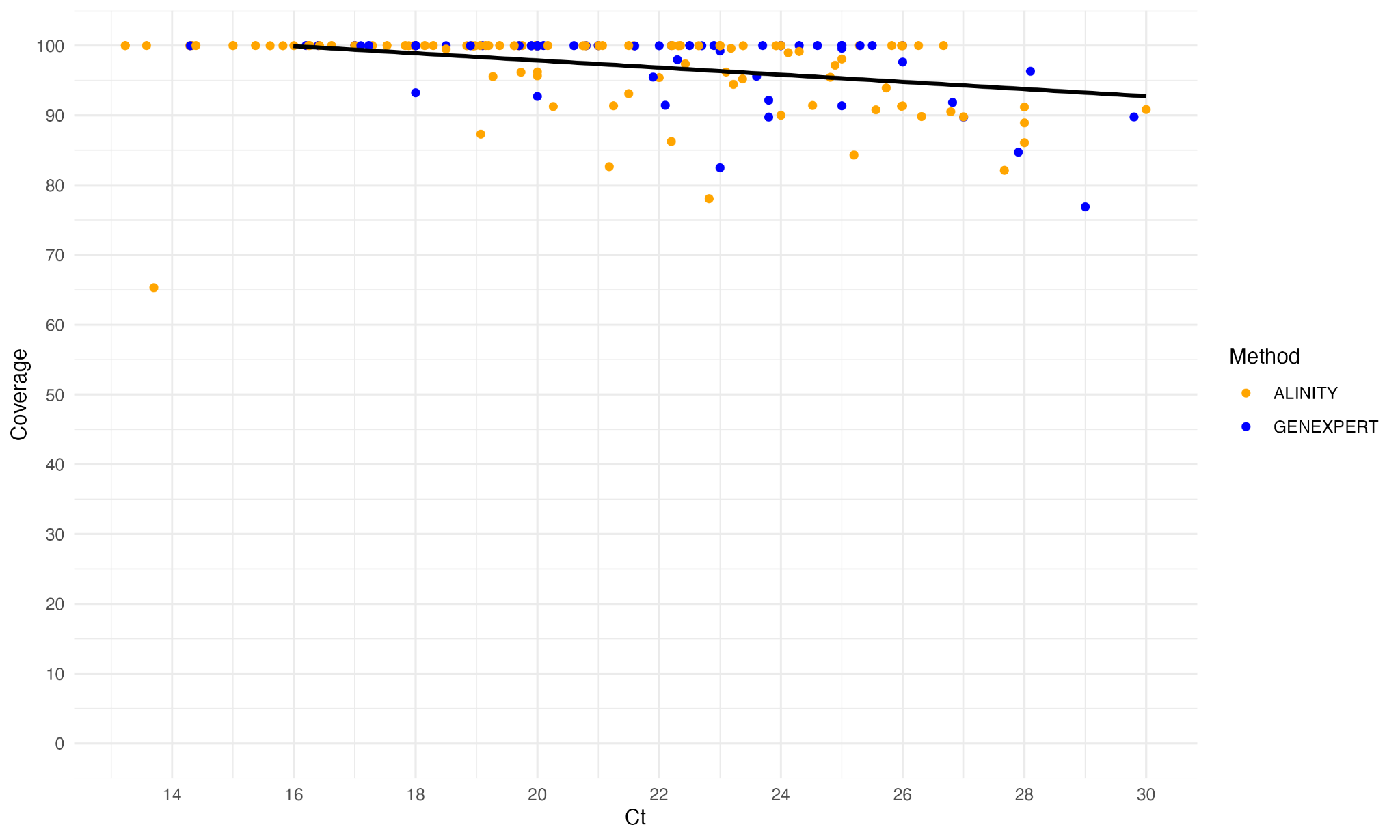

**Figure S2.** Correlation analysis of the breadth of coverage and the cycle threshold (Ct) in RSV samples sequenced with Oxford Nanopore Technologies (*p*=2.81x10^-8^, rho= -0.405).

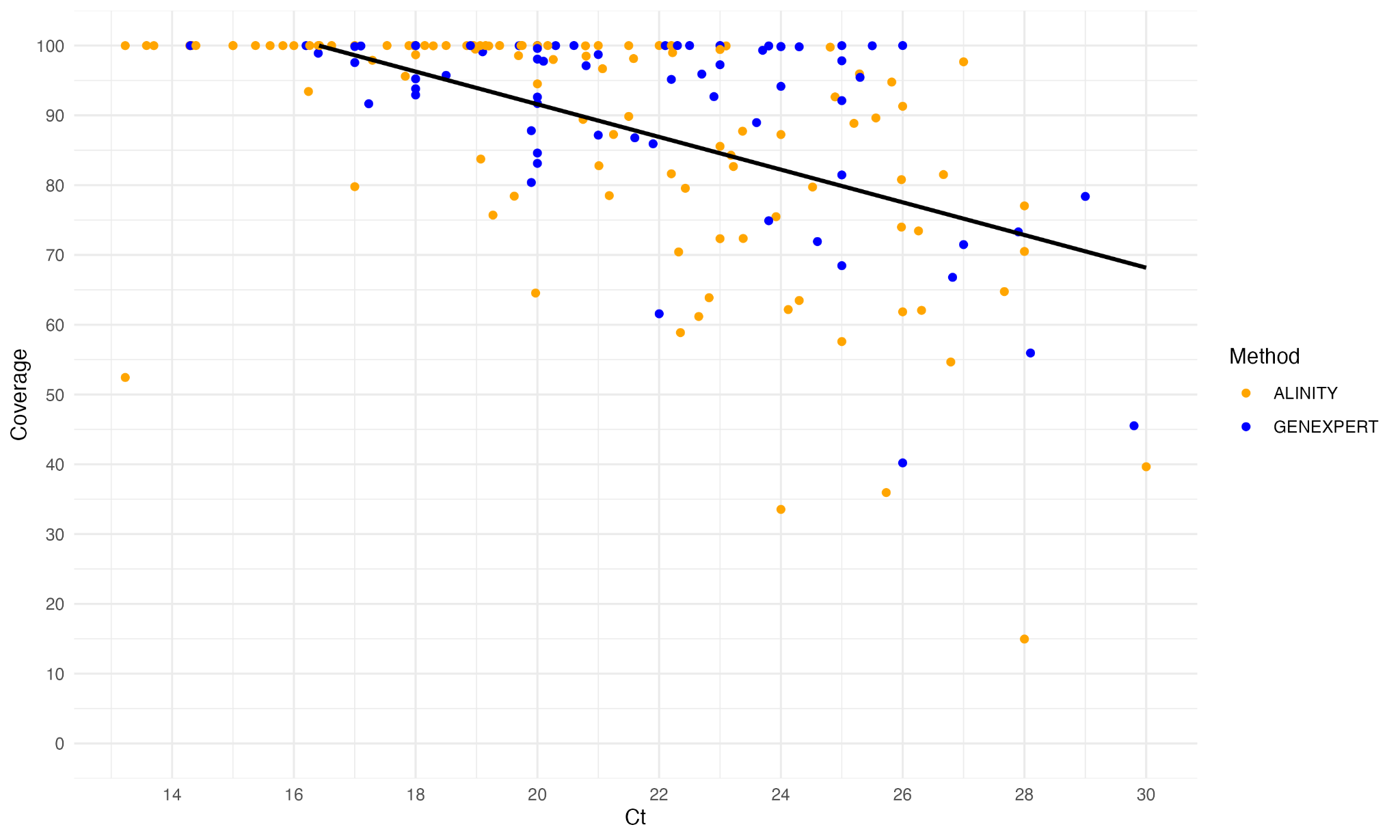

**Figure S3.** Correlation analysis of the breadth of coverage and the cycle threshold (Ct) in RSV samples sequenced with Illumina (*p*=1.56x10^-17^, rho= -0.586).

**Table S1.** Frequency of each RSV lineage assigned by Nextclade.

| **Clade** | **Oxford Nanopore Tech. (n=175)** | **Illumina (n=175)** |
| --- | --- | --- |
| A.D.1 | 12 | 12 |
| A.D.1.5 | 31 | 31 |
| A.D.1.8 | 1 | 2 |
| A.D.3 | 3 | 3 |
| A.D.3.1 | 3 | 3 |
| A.D.4 | 11 | 11 |
| A.D.5.1 | 4 | 4 |
| A.D.5.2 | 20 | 21 |
| A.D.5.3 | 2 | 2 |
| B.D.4.1.1 | 5 | 5 |
| B.D.E.1 | 83 | 81 |

**Table S2.** Demographic data of the patients with symptoms compatible with a viral infection included in the study (n=173).

| **Variable** | | **Count, n (%)** |
| --- | --- | --- |
| **Sex, male** |  | 83 (47.98) |
| **Age interval, years** | <4 | 84 (48.56) |
|  | 4-5 | 3 (1.73) |
|  | 6-60 | 20 (11.56) |
|  | >60 | 66 (38.15) |
| **Patient status** | Hospitalized | 27 (15.61) |
|  | Non hospitalized^$^ | 146 (84.39) |

^$^Patients admitted to the emergency room or outpatients.
